# Adapting models with single time-to-event outcomes to include a competing outcome: an exemplar adjusting risk of recurrence after nephrectomy for clear cell renal cell carcinoma for death from other causes

**DOI:** 10.1101/2025.05.22.25328132

**Authors:** Georgia Stimpson, Juliet A Usher Smith, Grant D Stewart, Paul Pharoah, Hannah Harrison

**Affiliations:** Department of Public Health and Primary Care, University of Cambridge, Cambridge, UK; CRUK Cambridge Centre, University of Cambridge, Cambridge, UK; Department of Surgery, University of Cambridge, Cambridge, UK; Department of Computational Biomedicine, Cedars-Sinai Medical Centre, Los Angeles, CA, USA

**Keywords:** Competing risks, Kidney Cancer, renal cell carcinoma, RCC

## Abstract

**Background:** Risk prediction models, in particular prognostic models, are used by clinicians to inform care and communicate risks to patients. However, many time-to-event models typically consider only one disease-specific outcome, which leads to overestimation of risk in populations where other-cause mortality is high. An example of this is the widely used Leibovich model, which models distant metastatic recurrence risk in patients with clear cell renal cell carcinoma (ccRCC, the most common form of kidney cancer) who have been treated surgically with radical nephrectomy.

**Methods:** In this study, we describe a novel approach for adapting existing risk prediction models retrospectively to include adjustment for a competing outcome, using population level data. We apply this approach to the Leibovich model, using life tables from the OZice of National Statistics, to generate the Leibovich Plus model and then illustrate the impact of increasing age on estimated risk of recurrence using both models.

**Results:** Comparing the predicted risk from the Leibovich model with the predicted risk of distant metastatic recurrence using the Leibovich Plus model, we show how distant-metastatic recurrence risk is overestimated when competing risks are not considered, particularly in older patients with high-risk tumours when using only a disease-specific outcome. For example, the risk of distant metastatic recurrence in individuals with a high-risk tumour pathology is 84.6% in a 55 year old individual after 10 years, but drops to 52.1% in an 85 year old individual with the same tumour pathology after 10 years.

**Conclusions:** This work describes an approach for adapting existing time-to-event models with disease-specific outcomes to include a competing outcome without the need for new data and illustrates the impact incorporation of competing risks has on estimated risk, particularly in older populations with high overall mortality risk. Such models, for example, the Leibovich Plus model for RCC, can be used in clinical consultations to provide a risk of recurrence adjusted for the risk of death from other causes.

## 1 Background

Risk prediction models utilise survival modelling techniques to generate an individual’s likelihood of an outcome, such as disease onset, disease recurrence or disease-specific survival. Risk models are particularly common in the cardiovascular(1) and oncological fields(2–4), where models such as QRISK-3(5) and BOADICEA(6) are well-established within clinical care. Prognostic models form a subset of risk models and predict an outcome of interest (often recurrence or cancer-specific survival) in a cohort already diagnosed with the disease of interest (such as PREDICT Prostate(7)). The target population for these models is also often at an increased risk of other-cause mortality due to shared risk factors such as age, comorbid diseases such as hypertension, or lifestyle factors. However, in many disease-specific survival models, individuals who die of other causes are censored in the same way as patients who reach the end of follow-up. As such, the nature of the competing risk, and the relative importance of the disease-specific risk, is lost. This is especially relevant when considering populations with a high proportion of frail or elderly individuals(8).

There are many methods for developing competing risks models. One of the most popular, the Fine & Gray model(9), utilises a proportional hazards approach using the cumulative incidence function. Here, the same risk variables are used to model both the outcome of interest and the competing outcome. This can increase the risk of overfitting as variables may be associated with one outcome only. An alternative is to use a joint modelling approach, whereby the risk factors for a given disease-specific outcome and a competing outcome can be identified and included into two models separately(7,10). These two submodels can then be combined to estimate overall risk, and the estimates for two outcomes reweighted with respect to this overall risk. Historically this method has been used at the model development stage, where a new model for the outcome of interest is generated using a dataset containing information about important risk factors, the outcome of interest and the competing outcome. The reception and adoption of new competing risk models therefore requires the disregarding of previous prognostic models for disease-specific outcomes that may already be widely accepted by the clinical community.

In this paper, we describe an alternative approach in which the disease-specific risk is reweighted with respect to a population-level outcome (such as other-cause mortality risk), calculated from existing population-level datasets. We use the recurrence of clear cell renal cell carcinoma (ccRCC), as an exemplar. Kidney cancer, of which the majority are renal cell carcinoma (RCC) is the 6^th^ most common cancer in the UK(11), with 80% of cases being clear cell type (12). Most cases of ccRCC are treated surgically, historically radical nephrectomy (full kidney removal), with partial nephrectomy and ablation becoming more common in recent years(12). More than 60% of kidney cancer diagnoses occur in the over 65s(13) and around 35% of kidney cancer patients are multimorbid at first diagnosis(14). The use of risk prediction models after nephrectomy to inform surveillance is recommended within clinical guidelines(15).

One of the most widely used models is the Leibovich score (16–19). It has been shown to accurately predict the risk of both recurrence and death from kidney cancer in multiple external validations(20). The Leibovich model was developed using data on time to distant metastatic recurrence, right censored at the last follow up or death(19), in a cohort of adult patients who had undergone radical nephrectomies for non-metastatic ccRCC at the Mayo Clinic, USA between 1970 and 2000.

With this work, the aims are to 1) describe how well-established, univariate outcome, clinical risk prediction models can be adapted for competing risks frameworks using population-level risk outcomes, 2) provide a use case for this approach in the real world, using the Leibovich model as an exemplar.

## 2 Methods

### 2.1 Competing Risk Methodology

#### 2.1.1 General Case

Given a Cox Proportional Hazards model, a time dependent cumulative hazard for a disease-specific outcome (*H*_*D*_(*t*|*X*_*D*_)) can be described as in Equation 1, where *H*_*D*0_(*t*) is the baseline cumulative hazard at time *t*, β represents the coefficient of each included risk factor, and *X*_*D*_ the individual’s characteristics. For example, the cumulative hazard of ccRCC recurrence at time t after surgery, where *X*_*D*_ includes tumour-specific variables such as stage or grade.

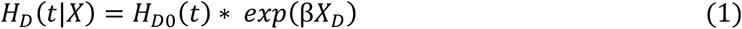

From this, the survival function at time t, *S*_*D*_(*t*|*X*_*D*_), (Equation 2), and the risk function at time t, *R*_*D*_(*t*|*X*_*D*_) (Equation 3) can be calculated.

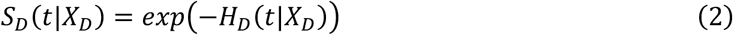

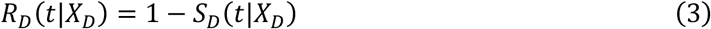

It is possible to generate the absolute risk of a competing population-level outcome at time *t* with respect to some demographic factors *X*_*P*_, *R*_*P*_(*t*|*X*_*P*_), using an independent population-level dataset. For example, the age and sex matched risk of mortality in the UK can be taken from the National life tables provided by the Office for National Statistics (ONS) for the year 2024(21).

Then, using established methods for the joint modelling of competing risks(7,10), *R*_*D*_(*t*|*X*_*D*_) and *R*_*P*_(*t*|*X*_*P*_) can be combined to generate the cumulative risk of both outcomes *R*_$_(*t*|*X*) (in our example, kidney cancer recurrence and death from other causes) at time *t* with respect to an individual’s characteristics (*X*), as given in Equation (4).

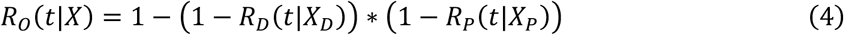

The combined risk of both outcomes can then be reweighted by the relative hazard of the disease specific and population-level competing outcome, to produce the relative cumulative disease-specific risk (*CR*_*D*_(*t*|*X*)) and the relative cumulative population-level competing risk (C*R*_*P*_(*t*|*X*)) as in Equations 5.1 and 5.2. In our example, this gives the risk of kidney cancer recurrence adjusted for the age and sex specific risk of death from other causes (and vice versa), as below:

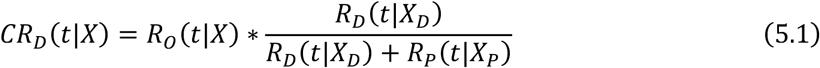

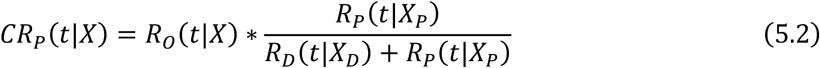

#### 2.1.2 Application to Kidney Cancer Prognosis

The Leibovich model is a Cox proportional hazards model for censored time to distant metastatic recurrence in patients treated with nephrectomy for ccRCC(19). The model includes pathological tumour stage, regional lymph node status, tumour size, nuclear grade, and tumour necrosis status. Using the coefficients from the Leibovich model, the Leibovich score was developed to further contextualise an individual’s risk. This gives an integer score ranging from 0 (a pT1a tumour less than 10cm in size, with a nuclear grade of 1, no lymph involvement, and no necrosis) and 11 (a pT4a tumour bigger than 10cm in size, with a nuclear grade of 4, lymph involvement and necrosis). There are also three broader categories which are widely used to stratify individuals for follow-up after surgery(22), based on the similarity of the risk profiles of the Leibovich score: low risk (Leibovich score 0-2), intermediate risk (Leibovich score 3-5) and high risk (Leibovich score 6-11).

To calculate the absolute tumour-specific distant metastatic recurrence risk, the authors of the paper describing the development of the Leibovich model(19) were contacted, and the baseline hazards were provided. The coefficients of the risk factors used in the model were extracted from Table 3 of Leibovich, B. C. et al. 2003(19). These were then used to calculate *H*_*D*_(*t*|*X*_*D*_), the cumulative hazard of distant metastatic recurrence with respect to individual’s tumour characteristics (*X*_*D*_). The age and sex-specific population-level estimates of absolute risk of death at time *t* (*R*_*P*_1*t*|*X*_*a*&*e*,*sex*_3) were generated using the Office of National Statistics (ONS) projected period life tables from 2024(21). Then equations (4), (5.1) and (5.2) were then used to generate the cumulative risk of distant metastatic recurrence (*CR*_*D*_(*t*|*X*)) and the cumulative risk of death (C*R*_*P*_(*t*|*X*)). We then describe this outcome as the Leibovich Plus outcome (i.e. Leibovich model plus the risk of death).

### 2.2 Assumptions

#### 2.2.1 General Assumptions

For this reweighting of models to incorporate a competing outcome to be appropriate, it is necessary to make the following assumptions. First, we must assume that the disease-specific outcome is not observed, or observed in such a small subset of cases, to make the impact of this outcome on the dataset used to model the competing population-level outcome negligible. In our example, this means assuming that cases of kidney cancer recurrence are not associated with a significant proportion of deaths observed in the UK general population.

Second, if using population level data for the competing outcome, we must assume that a disease-specific cohort would have an equivalent risk of the competing outcome when the disease-specific outcome is ignored. In our example, this means assuming that individuals diagnosed with kidney cancer have the same risk of dying from other causes as men and women of the same age in the general UK population.

### 2.2 Validity of Assumptions for ccRCC and the Leibovich Model

For the Leibovich Plus outcome, our assumption was that the proportion of patients who die of kidney cancer in the general UK population is small (such that the inclusion of kidney cancer deaths in the population level estimates of the risk of death is trivial and does not substantially contribute to the probability of dying) is reasonable. There were 4,700 deaths annually in the UK between 2017 and 2019 due to kidney cancer(13), compared to approximately 610,000 deaths annually in the same period in the UK(23). We should also consider that patients may go on to die after recurrence (from either ccRCC or other causes), leading to the same patient being double counted, however the number of cases where this occurs is small compared to the general UK population.

For the Leibovich Plus outcome, we further assume that those with ccRCC who have been treated with a radical nephrectomy do not differ from the UK general population when age and sex matching is considered. In practice this is unlikely to be the case, as there are lifestyle factors such as smoking(24) and obesity(25), and comorbidities such as hypertension(26) which are associated with an increased risk of ccRCC and other cause mortality. However, it is not possible to model these features without a prospective study (in which the relative risk of recurrence and mortality is calculated in the same cohort) and is a limitation of our approach.

### 2.3 Risk Classifications

We computed the estimated risks from the Leibovich and Leibovich Plus models using three hypothetical tumour phenotypes. The “Low Risk” tumour has a pathological T stage of 1a, no regional lymph node involvement, a size of less 10cm, a nuclear Grade of 1 and no necrosis (Leibovich score of 0). The “Intermediate Risk” tumour has a pathological T stage of 2, no regional lymph node involvement, a size of less 10cm, a nuclear Grade of 3 and no necrosis (Leibovich score or 3). The “High Risk” tumour has a pathological T stage of 3b, regional lymph node involvement, a size of greater than 10cm, a nuclear Grade of 3 and no necrosis (Leibovich score of 8).

## 3 Results

An overview of the Leibovich Plus predicted risks for the three “Low Risk”, “Intermediate Risk” and “High Risk” tumour phenotypes in men aged 55, 65 and 75 are given in Table 1, and the predicted risks for all combinations of age, sex and tumour features are given in Supplementary Table 1.

**Table 1:** Leibovich Plus predictions by tumour phenotype and age.

| Tumour Phenotype | Model | Age (years) | Sex | Risk of Distant Metastatic Recurrence by Follow-Up (%) |  |  | Other Cause Death by Follow-Up (%) |  |  | Metastases-Free Survival No Recurrence by Follow-Up (%) |  |  |
| --- | --- | --- | --- | --- | --- | --- | --- | --- | --- | --- | --- | --- |
|  |  |  |  | 1 year | 5 year | 10 year | 1 year | 5 year | 10 year | 1 year | 5 year | 10 year |
| Low Risk | Leibovich |  |  | 1.17 | 2.98 | 4.33 |  |  |  | 98.83 | 97.02 | 95.67 |
|  | Leibovich Plus | 55 | M | 1.17 | 2.94 | 4.22 | 0.00 | 2.96 | 6.81 | 98.83 | 94.11 | 88.97 |
|  |  | 65 | M | 1.17 | 2.92 | 4.19 | 0.99 | 6.85 | 15.45 | 97.84 | 90.23 | 80.36 |
|  |  | 75 | M | 1.16 | 2.91 | 4.17 | 2.97 | 16.57 | 38.44 | 95.86 | 80.53 | 57.40 |
|  |  | 85 | M | 1.16 | 2.90 | 4.16 | 1.16 | 2.90 | 4.16 | 89.93 | 52.39 | 19.13 |
| Intermediate Risk | Leibovich |  |  | 10.99 | 25.79 | 35.39 |  |  |  | 89.01 | 74.21 | 64.61 |
|  | Leibovich Plus | 55 | M | 10.99 | 25.09 | 33.32 | 0.00 | 2.92 | 6.59 | 89.01 | 71.99 | 60.09 |
|  |  | 65 | M | 10.89 | 24.37 | 31.49 | 0.99 | 6.61 | 14.24 | 88.12 | 69.02 | 54.28 |
|  |  | 75 | M | 10.73 | 23.15 | 28.74 | 2.93 | 15.26 | 32.49 | 86.34 | 61.60 | 38.77 |
|  |  | 85 | M | 10.45 | 21.53 | 26.70 | 8.55 | 38.40 | 60.37 | 81.00 | 40.07 | 12.92 |
| High Risk | Leibovich |  |  | 46.56 | 79.91 | 90.47 |  |  |  | 53.44 | 20.09 | 9.53 |
|  | Leibovich Plus | 55 | M | 46.56 | 77.60 | 84.59 | 0.00 | 2.91 | 6.55 | 53.44 | 19.49 | 8.87 |
|  |  | 65 | M | 46.10 | 74.77 | 78.17 | 0.99 | 6.55 | 13.82 | 52.91 | 18.68 | 8.01 |
|  |  | 75 | M | 45.25 | 68.71 | 65.37 | 2.92 | 14.62 | 28.91 | 51.84 | 16.67 | 5.72 |
|  |  | 85 | M | 43.05 | 56.58 | 52.06 | 8.32 | 32.57 | 46.04 | 48.63 | 10.85 | 1.91 |

In Figure 1, we display the risk of distant metastatic recurrence generated by the original Leibovich model (black, dashed) and show how the risk of recurrence for the three tumour phenotypes changes depending on an individual’s age at surgery when using the Leibovich Plus model (coloured). The largest changes in risk are seen for older individuals and individuals with higher risk tumours.

**Figure 1:**
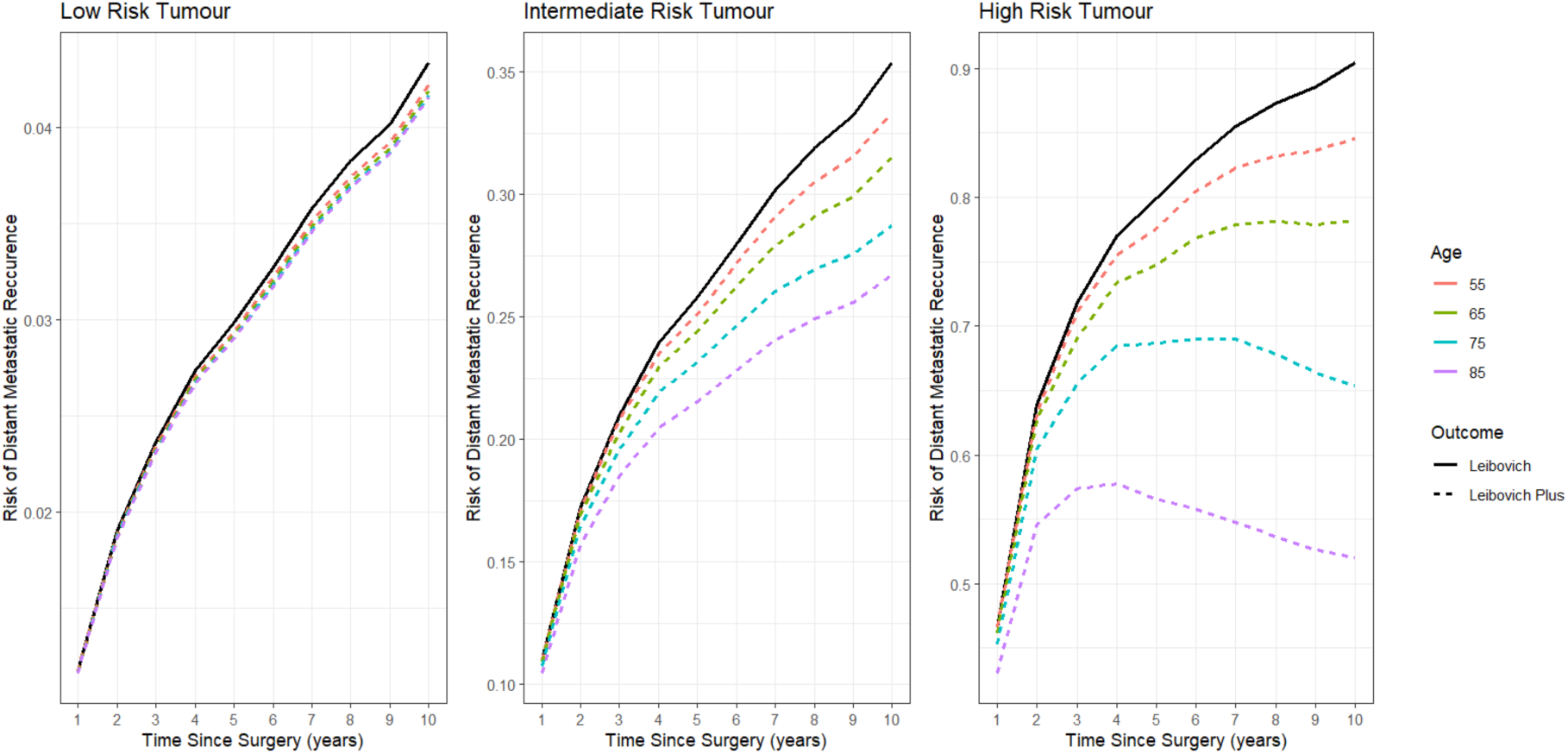
Impact of Leibovich Plus competing risks adjustment on the Leibovich Metastatic Recurrence Predictions

In Figure 2, for the same three tumour phenotypes, we show the competing risk of metastatic-recurrence (dark blue), other-cause death (grey) and metastases-free survival (green) for a man aged 55, 65, 75 and 85 years respectively. In the low risk tumour phenotype, the decrease in distant metastatic-recurrence risk as age increases is small (4.22% and 4.17% at 10 years in the 55 and 75 year old individuals, respectively), despite the increase in the risk of death from other causes (6.8% and 38.4% at 10 years in the 55 and 75 year old individuals, respectively). Therefore, the unadjusted Leibovich model provides a reasonable estimate of recurrence risk in this scenario. In the intermediate risk and high risk tumour phenotypes, however, the adjusted risk of ccRCC recurrence substantially decreases with age in the Leibovich-plus model, with the original Leibovich model shown to substantially overestimate metastatic-recurrence risk relative to other-cause death. For example, 10-years after surgery, the risk of distant metastatic recurrence in individuals with a high risk tumour is 84.6% in a 55 year old individual, but drops to 52.1% in an 85 year old individual after 10 years with the same tumour pathology. This overestimation of recurrence risk when considering only the disease specific outcome is particularly noticeable in individuals with high-risk tumours ages 75 and 85. In these individuals, the Leibovich Plus outcome predicts they have very low rates of metastasis-free survival 10 years after surgery (5.7% and 1.9% at 10 years in the 75 and 85 year old individuals, respectively).

**Figure 2:**
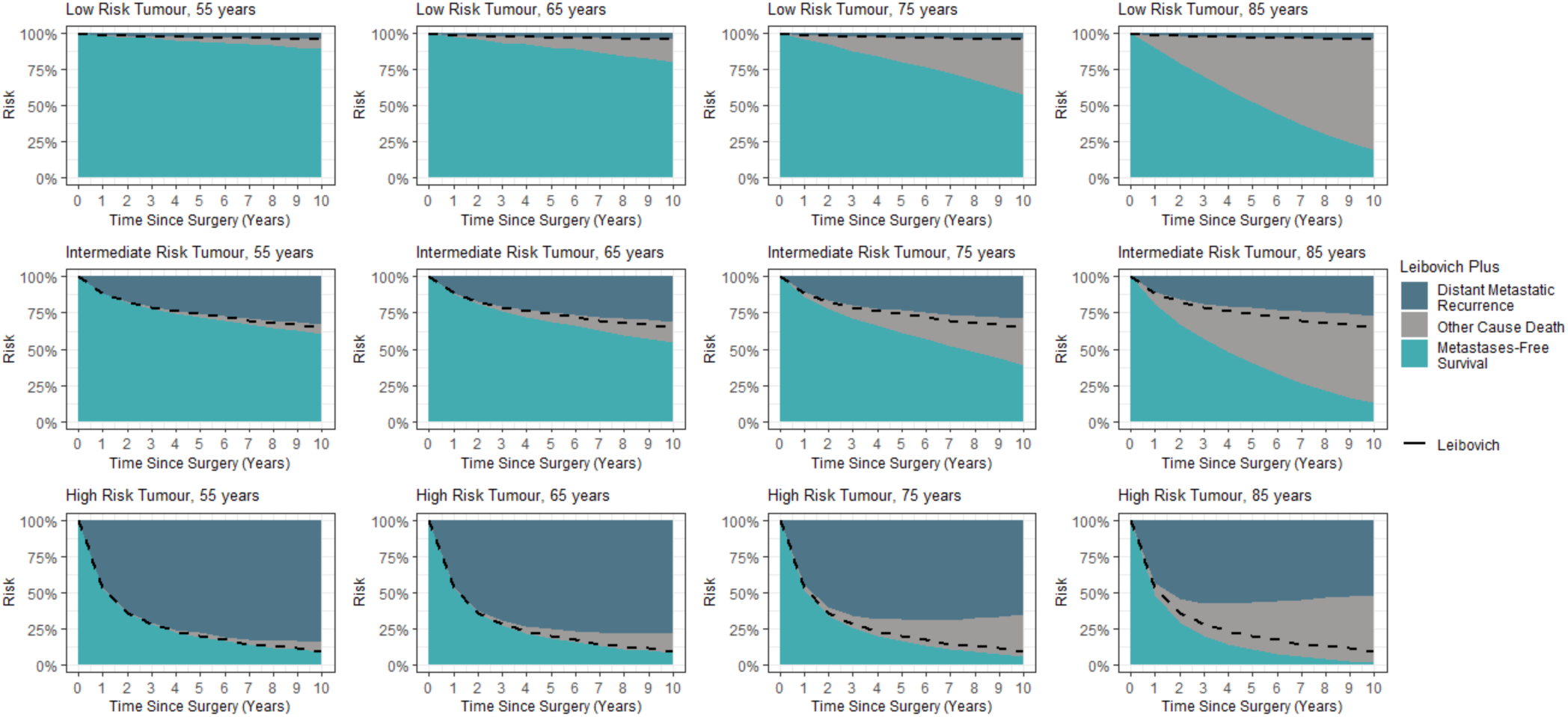
Distant Metastatic Recurrence, Other Cause Mortality and Metastases-Free Survival Risk by tumour and demographic phenotype

## 4 Discussion

There are many risk prediction models for predicting disease-specific onset, recurrence, and mortality using survival analysis that do not account for competing risks. However, predictions generated by these approaches are inaccurate in cohorts with a high likelihood of a competing outcome (such as high overall mortality), with the degree of overestimation directly linked to the patient-specific risk. In this paper we describe an approach that can be applied retrospectively to reweight risk predictions from existing survival models that consider only a single disease-specific outcome with respect to other-cause mortality. We apply this approach to the Leibovich model, which is widely used to predict kidney cancer recurrence, updating that model using UK population wide age and sex specific mortality data to develop a model that accounts for other-cause mortality, the Leibovich Plus model. Comparing the predictions generated by these two models, we show how incorporating competing risks reduces the substantial overestimation that arises when considering only single disease-specific outcomes, particularly for older patients, or those with a “high risk” tumour phenotype.

A key benefit of this approach is that it can be performed retrospectively and using relevant population level data to model the competing outcome. Therefore, no new prospective data is required to construct the augmented outcome. It is worth noting, however, that an external validation of any new competing risks outcome (with a comparison to the performance of the original univariate risk model) would be required to determine the potential for application in a clinical setting.

A further limitation of this approach is that the original model development cohort and the population-level data used to model the competing outcome may not be directly comparable. For example, the Leibovich score was developed in a cohort who underwent surgery at the Mayo Clinic, Minnesota, between 1970 and 2000(19), and as such the censoring of patients due to death is likely to differ from the Office of National Statistics (UK) period life tables from 2024(21).

However, given that when creating the Leibovich Plus model we were intending to use this outcome to communicate the risk of recurrence (with adjustment for death from other causes) in urology clinics in the UK(27), the use of recent UK data for the outcome of other-cause mortality was appropriate. The approach could easily be repeated using different population-level data for other settings.

A common clinical application of risk models is to stratify the population of interest into groups for surveillance or intervention. The Leibovich score and risk classification system (low, intermediate and high) are typically used to communicate risk of cancer recurrence to patients and to select an appropriate surveillance schedule following kidney cancer surgery (with the aim of detecting recurrent disease)(15). The use of a competing risks framework, as described in this paper, not only provides more accurate estimates of recurrence but also enables clinicians and patients to consider risk of recurrence in the context of other-cause mortality and facilitate more individualised and nuanced discussions about surveillance strategies. However, it adds a layer of complexity to the selection of suitable risk groupings, as the risk estimates are not concordant over time. Using the Leibovich Plus model, an individual A, who has a higher risk of recurrence using the Leibovich Plus at 5 year follow up compared to individual B, may have a lower recurrence risk at 10 years if patient A is significantly older than patient B. Consequently, classification into risk groups needs to be considered carefully in each context, with either reclassification over time or clustering of the overall risk predictions with time considered. Consideration also needs to be given to how best to communicate these risks to patients, particularly in the context of a new diagnosis of disease and for older co-morbid patients where the risk of other-cause mortality will be high.

### 4.1 Conclusion

With this work, we describe an approach for adapting risk models of disease-specific outcomes to include an adjustment for a competing outcome of interest using population level data. Compared to univariate risk models, which overestimate the disease-specific outcome in specific subgroups of the population, this approach allows more nuanced conversations around care by presenting competing population-level outcomes, such as other cause mortality. We demonstrate this by augmenting the Leibovich model, which is widely used to predict prognosis following kidney cancer surgery, with population level data to generate the novel Leibovich Plus outcome. Further work is ongoing to validate this outcome, to understand how best to implement it within clinical practice and produce a communication tool to allow the calculation of individual risks (27).

## Data Availability

All data analysed in this paper are available online.

## Abbreviations

ccRCC: Clear cell RCC
ONS: OZice for National Statistics
RCC: Renal cell Carcinoma

## 6 Declarations

### 6.1 Ethics approval and consent to participate

Not applicable

### 6.2 Consent for publication

Not applicable

### 6.3 Availability of data and materials

The data supporting the conclusions of this article are included within the additional files of the article.

### 6.6 Authors’ contributions

GS analysed the data, produced the results, drafted the paper and substantially revised it. HH conceptualised the work, acquired the data, drafted the paper and substantially revised it. JUS and GDS conceptualised the work and substantially revised it. PP helped to design the analysis. All authors read and approved the final manuscript.

## 6.7 Acknowledgements

For this project, we would like to acknowledge the insights of the PREDICT Kidney workshop attendees, who, in part, motivated this work. We would also like to acknowledge Richard Jetten, a patient and co-applicant, and our patient panel: David Brownlees, Andrew Fear, Stewart Hoare, Lawrence Smith-Higgins and Tom Wallace. This project is hosted by the Cambridgeshire and Peterborough Integrated Care Board in partnership with the University of Cambridge. We would also like to acknowledge Bradley Leibovich and Christine Lohse from Mayo Clinic, Rochester, USA, and the team that developed the Leibovich model and provided us with the baseline hazards for the Leibovich score.

## 7 Lay Summary

Risk prediction models give patients and clinicians insights into how likely a specific outcome is to occur, such as a diseases onset, a disease recurring or death due to a given disease. In particular, prognostic models are used to look at one outcome in a group of patients with shared specific characteristics (such as with the same disease or on the same treatment). Traditionally, the methods used for these models only consider one outcome, and patients who experience a different outcome are “censored”. For example, in a model looking at recurrence of disease A, a number of years following surgery, those who die of disease B without a recurrence of disease A, say, 5 years after surgery are excluded from the model used to estimate the risk of recurrence at 6 years. Consequently, in groups of patients who have a high risk of death due to other diseases, traditional methods can overestimate risk.

In this paper, we describe a method for adapting pre-existing outcomes for the risk of death from other diseases. Key strengths of this work are that it can be performed on outcomes that are already well established in clinical care, and that it does not require new data, which can be difficult and time consuming to acquire.

We implement this method for an outcome called the Leibovich score, which models the risk of recurrence in some patients with kidney cancer who have been treated by having their kidney removed. We call our new outcome the Leibovich Plus outcome. We show that the Leibovich Plus risk is considerably lower than the Leibovich risk for patients who have cancers that are more likely to reoccur (e.g. bigger in size or more aggressive) or who are older. By describing the relative risk of the cancer returning and the risk of other-cause death, we aim to help clinicians have more informed conversations with patients about their prognosis and follow-up.

